# Correlation between daily infections and fatality rate due to Covid-19 in Germany

**DOI:** 10.1101/2020.08.03.20167304

**Authors:** Dieter Mergel

## Abstract

The officially reported daily Covid-19 fatality rate is modelled with a trend line based on a nominal day-to-day reproduction rate and a cosine to take account of weekly fluctuations. Although the time trajectories of officially reported infections and fatalities are pronouncedly different, the reproduction rates obtained therefrom are similar. The long-term effective reproduction rate is around 0.835 and the administrative measures to contain the pandemic seem not to have an immediate reducing effect but well the ease of restrictions an increasing one. The fatality trajectory represented by its trend line can be projected from the number of daily infections by assuming a time lapse between symptom onset and death between 17 and 19 days and a time-dependent nominal lethality. The time trajectory of this lethality increases from 2.5% at March 16 when public life was restricted to 6% within 20 days indicating relatively more infections of vulnerable people. After stipulating face mask wearing at April 27, the nominal lethality decreases down to 1% later in summer. A detailed analysis shows that mask wearing really reduces the number of fatal infections and the officially reported daily infections in May and June are less lethal than before.

## 1 Introduction

In an earlier paper, we have presented a mathematical procedure to lay a trend line through the reported number of daily infections in Germany, Sweden, and France based on a day-to-day reproduction rate and, if appropriate, a cosine to account for periodic weekly fluctuations (Mergel D 2020 1, Mergel D 2017). This procedure achieves effective noise reduction and allows to calculate an effective reproduction rate at the time of symptom onset of the infecting persons. It further allows to construct the infection time trajectory from the effective reproduction rate so that different progressions of the epidemic can be modelled. Hypothetically implementing part of the dynamics in Sweden onto that of Germany leads to about 9500 more fatalities up to the end of April, roughly estimated.

It has often been argued that the reported number of confirmed infections is not representative of the number of true infections due to peculiarities of the testing procedure so that the only reliable data are case fatalities.

In this paper, we:

- lay a trend line through the reported number of Covid-19 fatalities with the same procedure based on nominal reproduction rates,
- predict the fatality trajectory from the infection trajectory assuming a time-dependent lethality,
- check whether the reproduction and lethality trajectories are correlated to the administrative lock-down measures listed in Table 1.

**Table 1.** Lockdown measures of the German administration

|  |  |  |
| --- | --- | --- |
| $t =$ | | |
| 9 | 09.03. | mass events prohibited |
| 16 | 16.03. | public life restricted |
| 23 | 23.03. | private contacts officially restricted |
| 58 | 27.04. | face masks obligatory in public places |
| 107 | 15.06. | easing of lockdown |

Our approach is a pure data-scientific analysis of the numbers reported by the Robert-Koch-Institut. Epidemiological knowledge reported in the literature enters only for the definition of the effective reproduction rate and the time lapse between symptom onset and death. Nevertheless, we shall try to draw conclusions to be discussed under epidemiological aspects.

The nomenclature used in this paper is summarized in Table 2.

**Table 2.**
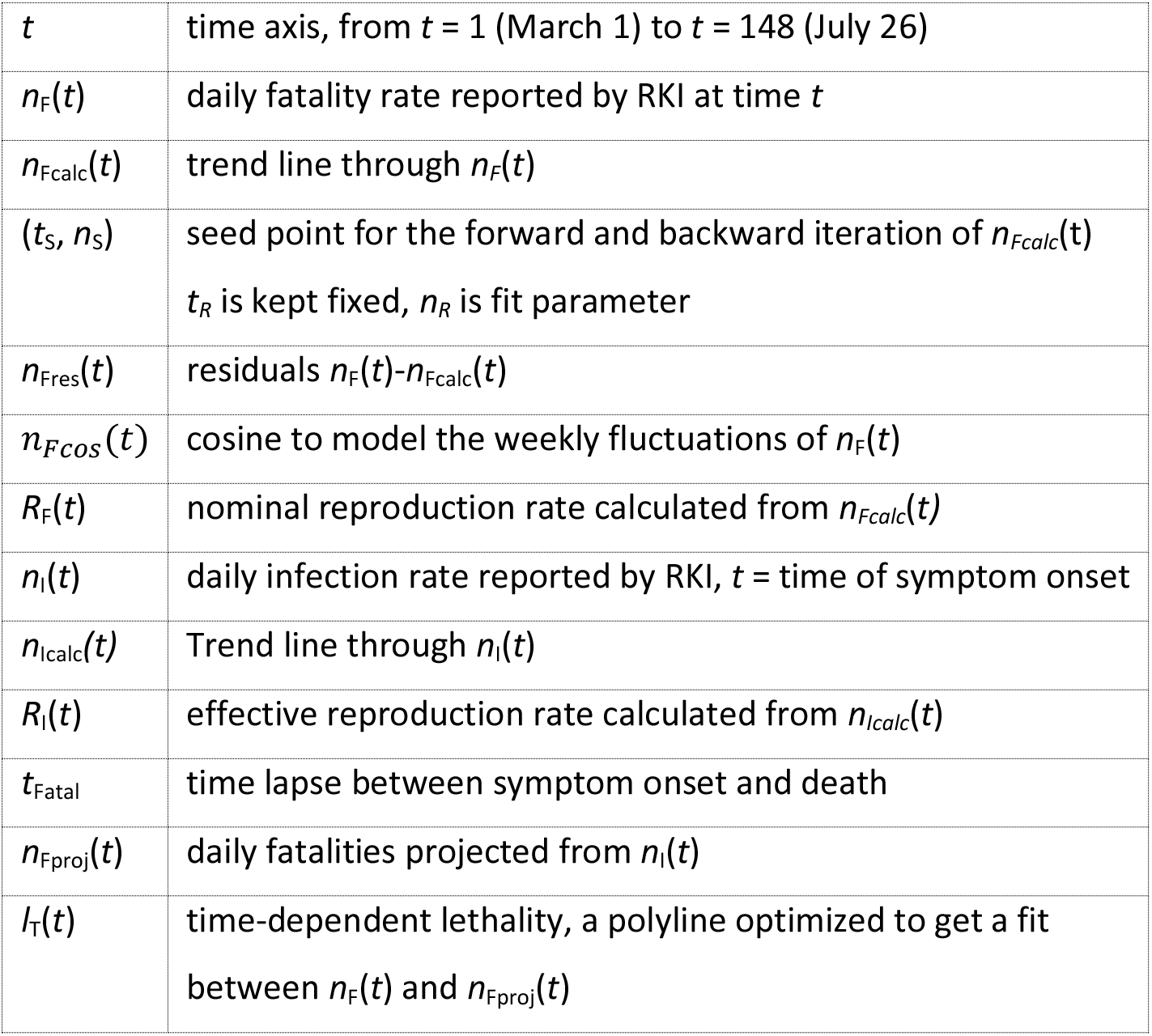
Data structure and nomenclature

## 2 Trend line for the daily fatality rate

Figure 1 shows the daily Covid-19 fatality rate *n*_F_ from march 9 (*t*=9) to July 26 (*t* = 148) as reported by the Robert-Koch-Institut (RKI 23-07-2020, 2) together with a trend line *n*_Fcalc_ modelled with a nominal day-to-day reproduction rate *r*_T_ of the type developed for the daily infections with Covid-19 (Mergel 2020, 1; Mergel 2017). The procedure to get the trend line is as follows:

**Figure 1.**
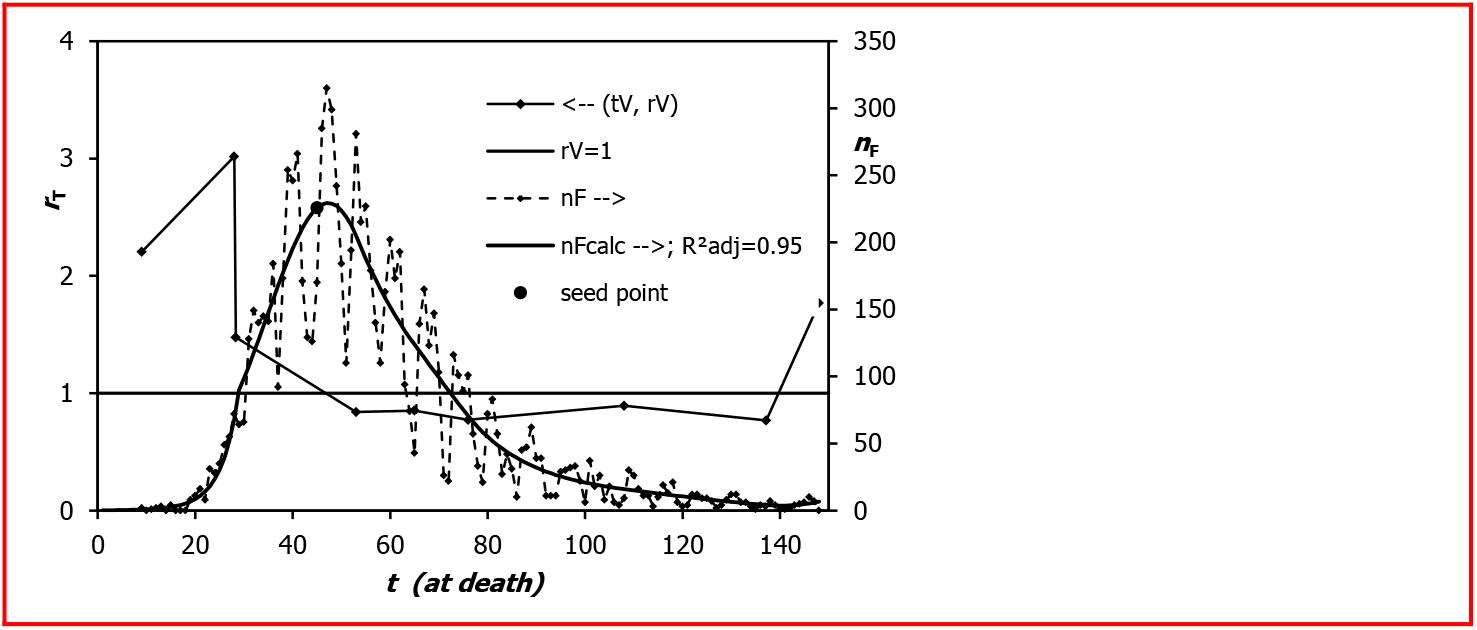
Daily fatality rate *n*_F_(*t*), trend line *n*_Fcalc_(*t*), and nominal day-to-day reproduction rate *r*_T_(*t*), *R*^2^_adj_ due to Equation 4 and Equation 5

A seed point (*t*_S_, *n*_S_) is chosen near the supposed maximum of the curve and the trend line is modelled, with *n*_Calc_(*t*_S_) = *n*_S_, for *t* ≥ *t*_S_ as:

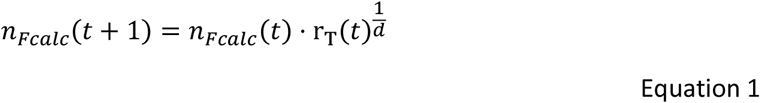

and for *t* < *t*_s_ as:

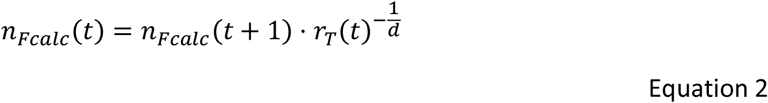

The power parameter *d* is set to 4, *d* = 4, because this is the most probable time span between the symptom onsets of the infect-*ing* and the infect-*ed* person (Böhmer M and others 2020) and *r*_T_ comes close to the effective reproduction rate. The position *t*_S_ of the seed point is kept fixed but the value *n*_S_ is a fit parameter.

The function *r*_T_*(t)* is modelled as a polyline with 9 vertices where the start and stop instants, *t* = 9 and *t* = 148 are kept constant and the other 16 parameters are treated as adjustable fit parameters.

Figure 2 displays the residuals *n_Fres_ (t) = n_F_(t) — n_Fcaic_(t)* of the fit of Figure 1 together with a trend line *n*_Fcos_ intended to represent weekly fluctuations:

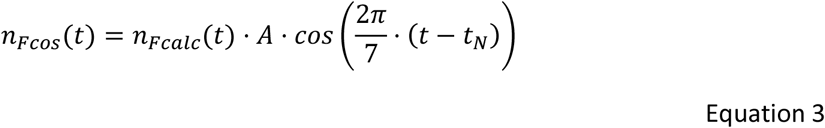

**Figure 2.**
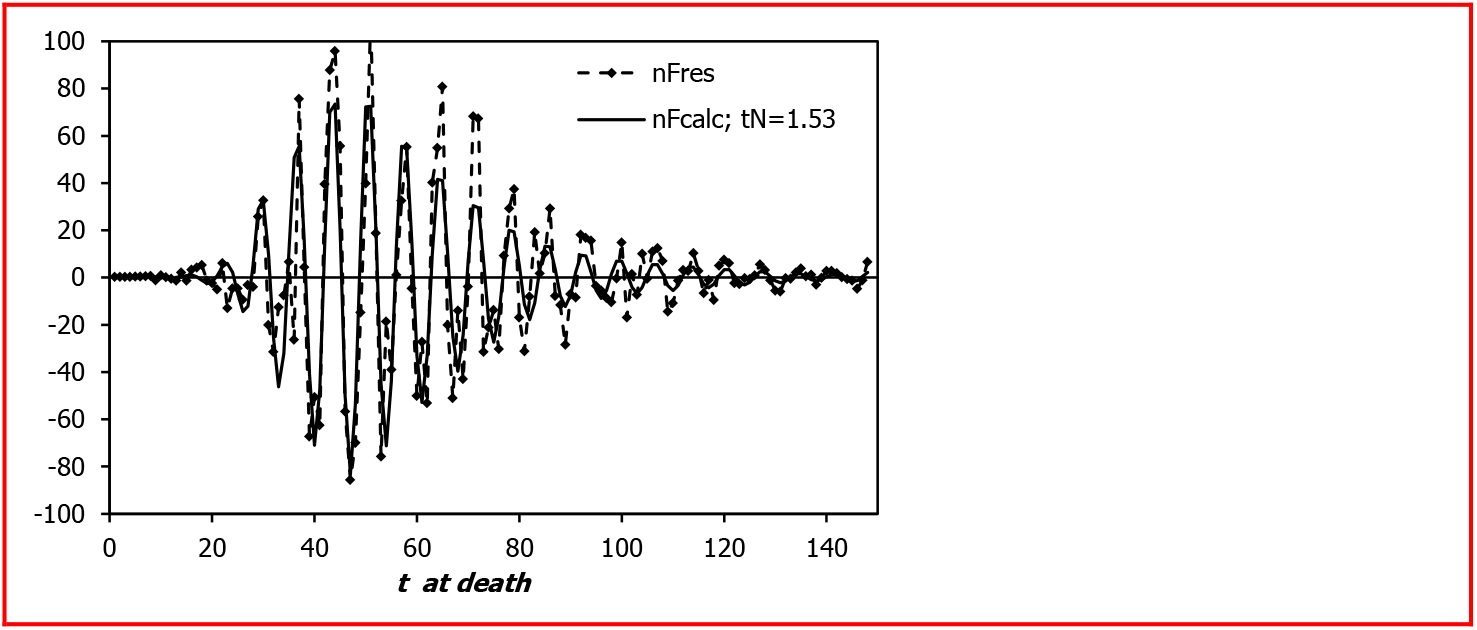
Residuals *n*_Fres_ of the fit of Figure 1 together with the cosine *n*_Fcos_ defined in Equation 3

The sum of the square deviations of the experimental *n*_Fres_ and the modelled *n*_Fcos_ residuals is taken as the target to be minimized with the 1 value of the seed point, the 16 parameters of the vertices and the 2 parameters *A* and *t*_N_ of the model residuals. So, the trend line in Figure 1 minimizes the sum of the square deviations of the residuals from the cosine model and therewith maximizes the adjusted coefficient of determination 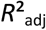 defined as

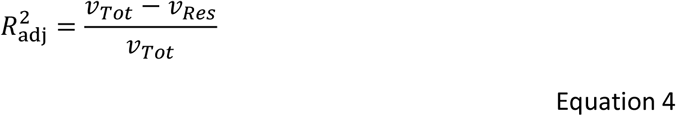

where *v*_Tot_ is the total variance of the *n*_F_ and *v*_Res_ the adjusted residual variance (adjusted for the decreased degree of freedom due to the number of fit parameters) calculated as:

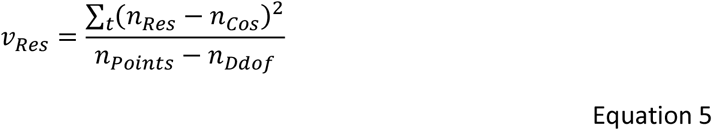

where *n*_Points_ is the number of data points (here 140) and *n*_Ddof_ the number of fit parameters (here 1 + 16 + 2 = 19). For the fit in Figure 1 and Figure 2, 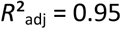 is achieved.

## 3 Projecting the fatality rate from the infection rate

For people dying of Covid-19, the time between symptom onset and death is reported to be 17, 18, 19 days in 90% of cases (Verity R 2020 and others). So, our model for projecting the fatality rate is:

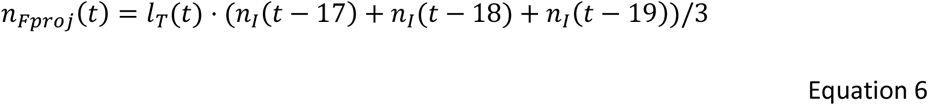

With *l*_T_ being a nominal lethality, called nominal because the reported number of infections is generally regarded to be systematically underestimated. Setting *l*_T_ = const. = 6% yields the projection *n*_Fproj_ displayed in Figure 3.

**Figure 3.**
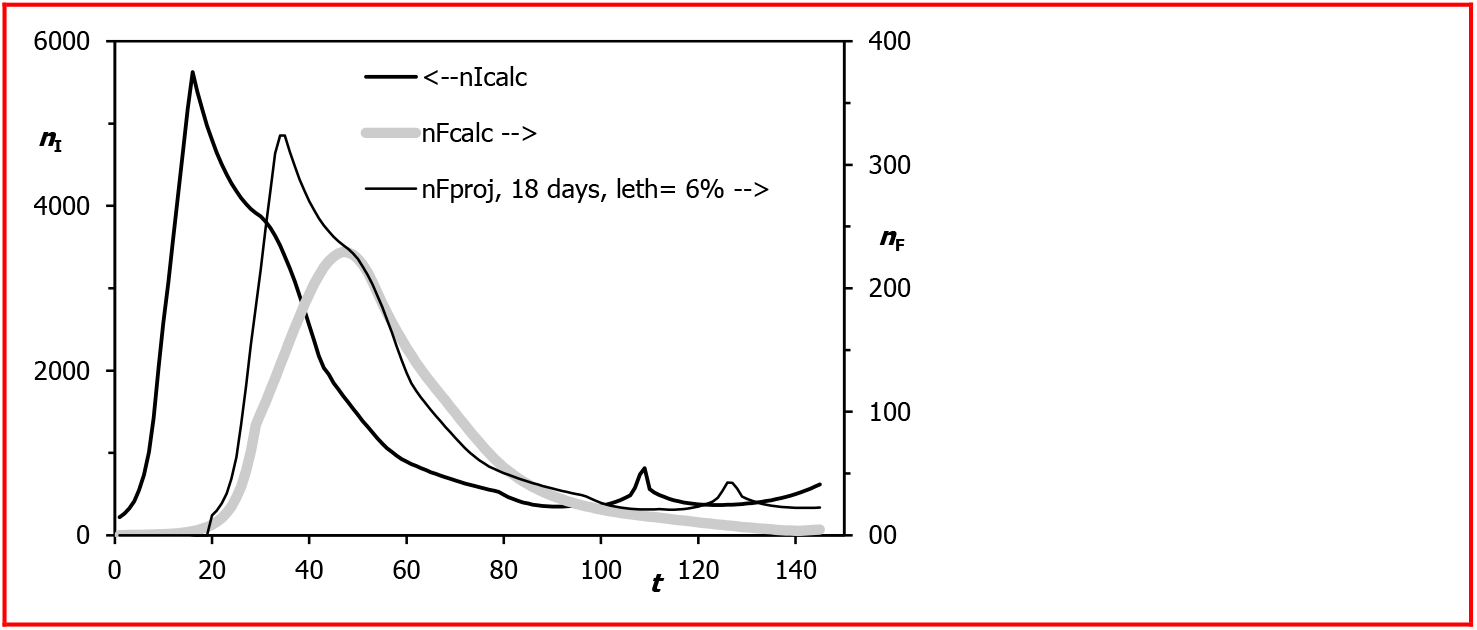
*n*_Fproj_ is the fatality rate projected from the trend line *n*_Icalc_ of the infection rate, here with constant lethality (6 %) and average time to death 18 days.

The time shift seems to fit with the onset of fatality and the approximately exponential decay after *t* = 50 so that the assumed average time of 18 days from symptom onset to death is compatible with the data. However, the characteristic shape of the fatality time trajectory is not captured. There seems to be a marked difference between an early and a later stage of the epidemic. We therefore try a second approach by modelling the lethality time-dependent, using again a polyline as for the infection rate and the fatality rate.

The result of the fit can be seen in Figure 4 where the *y* axes are logarithmic to check more easily the plausibility of the lethality trajectory that is approximately the ratio (or the distance in logarithmic plot) of the number of fatalities displayed over the presumed time of infection and the number of infections displayed over the time of symptom onset.

**Figure 4.**
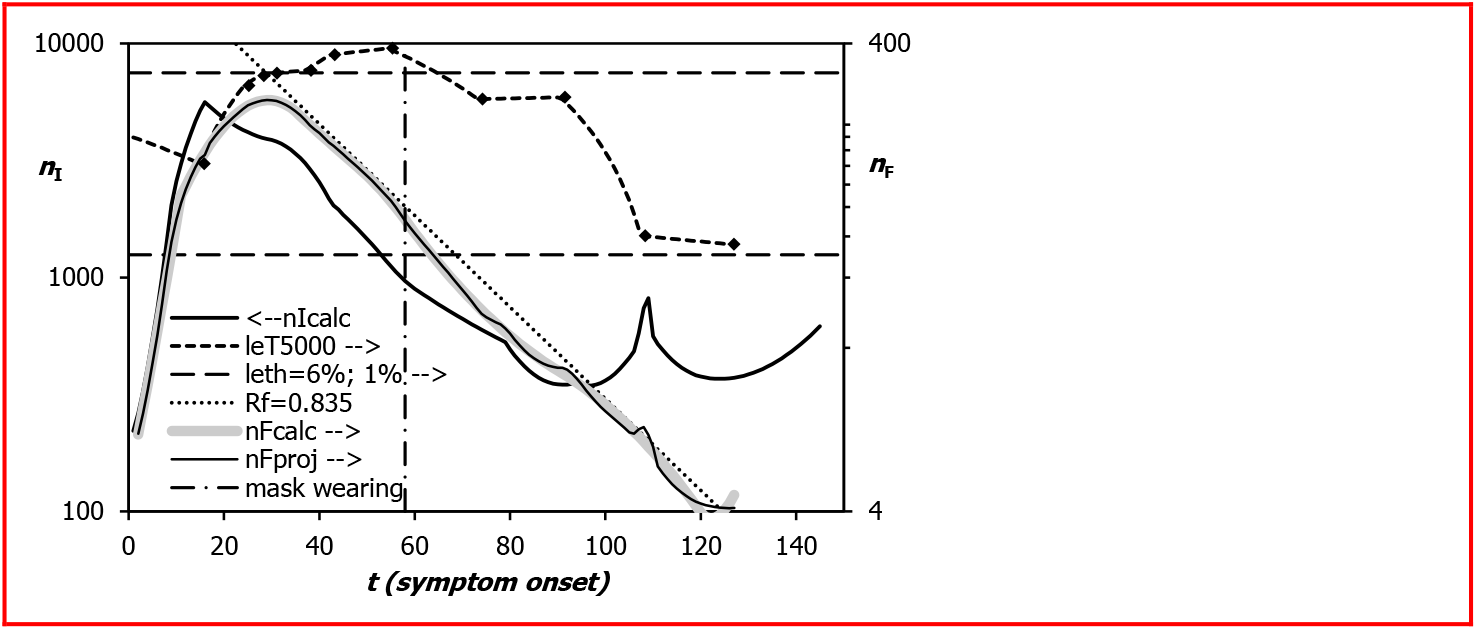
Semilogarithmic plot, fatality rate *n*_Fproj_ projected from the trend line *n*_Icalc_ of the infection rate with the time dependent lethality *le*_T5000_ as fit curve, “5000” means that the numbers have been multiplied by 5000 to fit to the secondary vertical axis

The polyline, designated *l*_T_, is defined by 12 vertices. It is displayed over the time of symptom onset, 18 days before death. Now, the daily fatalities, displayed over the time of symptom onset, are reasonably well modelled.

A similar fit is obtained when assuming an average time to death of 20 days leading to a better coincidence with the initial stage (as of *t* = 20) of the fatality rate. The two nominal lethality rates are displayed together in Figure 5 in linear scale. They represent the probability to die for a person with symptom onset at t. The vertical dashed lines indicate the administrative measures listed in Table 1 and intended to contain the spread of the virus.

**Figure 5.**
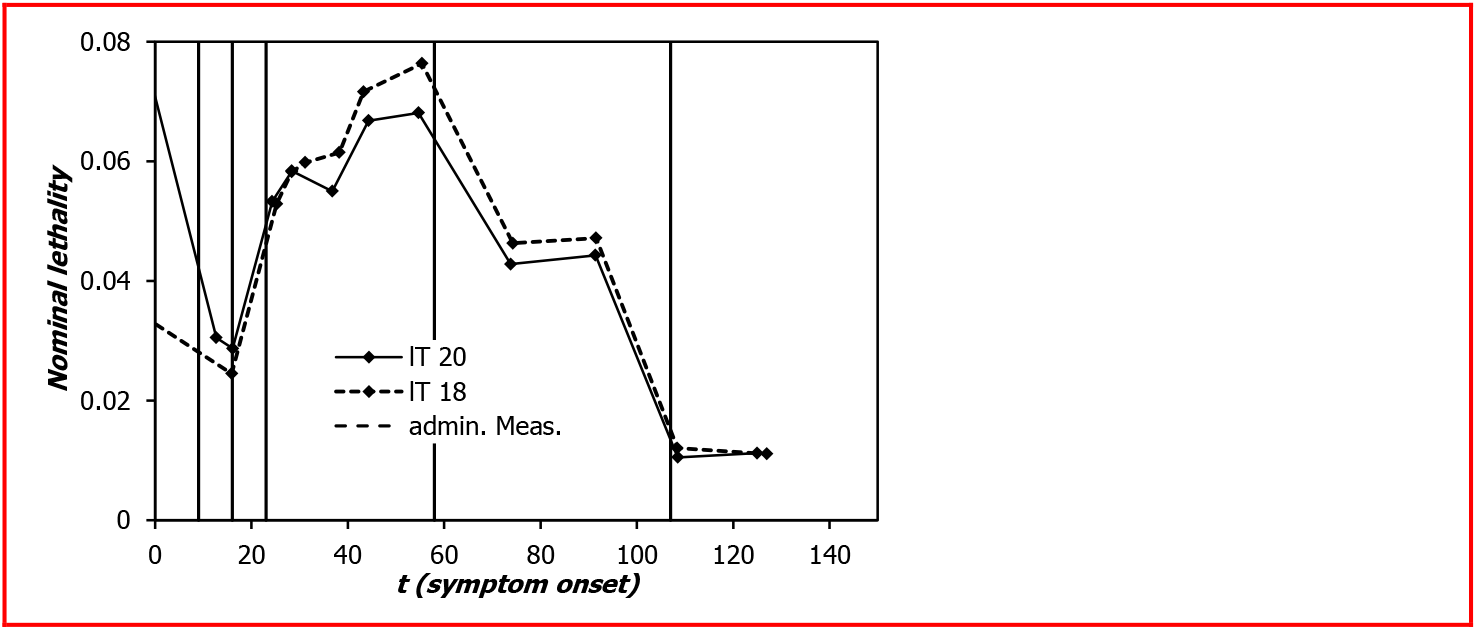
Nominal lethality trajectory for assumed average time to death alternatively 18 or 20 days

Their most prominent common features are:

- lethality dynamics changes for symptom onset at *t* = 16 (March 16) with rapid increase from 2.5% to above 6% for *t* > 30.
- nominal lethality decreases after *t* = 58 (April 27) down to about 1% at *t* = 125 (July 7), still higher than the Covid-19 fatality rate (0.1 to 0.6%) estimated from serological studies (Streeck H and others 2020, Bendavid and others 2020, Mergel 2020-2).

March 16 is the date when public life was restricted. The increase after that date may be because the infection has spread from younger and healthier mobile people being infected in superspreading leisure time events to more vulnerable persons.

April 27 is the date when mask wearing in shops and public transport became obligatory. The most surprising finding of this analysis is that stipulating mask wearing does not have a positive effect on the effective reproduction rate (see Figure 4) but is evidently correlated with the beginning of a decrease in nominal lethality.

Nominal lethality may decrease because:

- testing has captured more people with mild diseases;
- vulnerable population is better protected;
- Covid-19 lethality is really lower.

It is tempting to conjecture that mask wearing perhaps together with beneficial effects of spring and summer time causes really a decrease in Covid-19 lethality. A closer look at Figure 4 gives more clarity.

The dotted line in Figure 4 close to *n*_Fcalc_, the trend line through daily fatalities, represents an exponential decay *N·exp(a·t)* with *a = ln(0.835)/4* to indicate the theoretical course for *R*_f_ = 0.835. This line describes the long-term trend of *n*_Fcalc_ quite well. At *t* ≥ 58 we observe a decrease of daily fatalities below this line up to *t* = 95 (i.e. essentially in May), and a decreased decay of reported daily infections. So, the rapid decrease in nominal lethality, essentially the ratio of the two curves or the distance in logarithmic scale, is due to both effects. Nevertheless, we may conclude that mask wearing has the positive effect of reducing the number dangerous infections.

The discrepancy of the trajectories *n*_Icalc_ and *n*_Fcalc_ after *t* = 90 (i.e. in June) confirms that the officially reported daily infections are less lethal than before.

## 4 Reproduction rates

A nominal effective reproduction rate *R*_f_(*t*) is calculated from the trend line through the fatality trajectory as (Mergel 2020, 1):

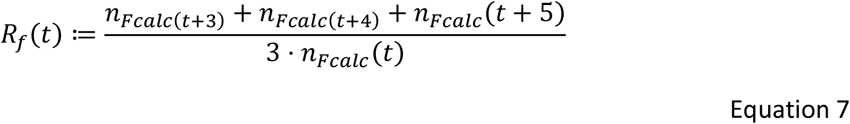

Figure 6 compares this reproduction rate *R*_f_*(t)* being shifted by *t*_Fatal_ = 18 days to earlier dates with the effective reproduction rate *R*_I_(*t*) obtained to model the daily number of infections.

**Figure 6.**
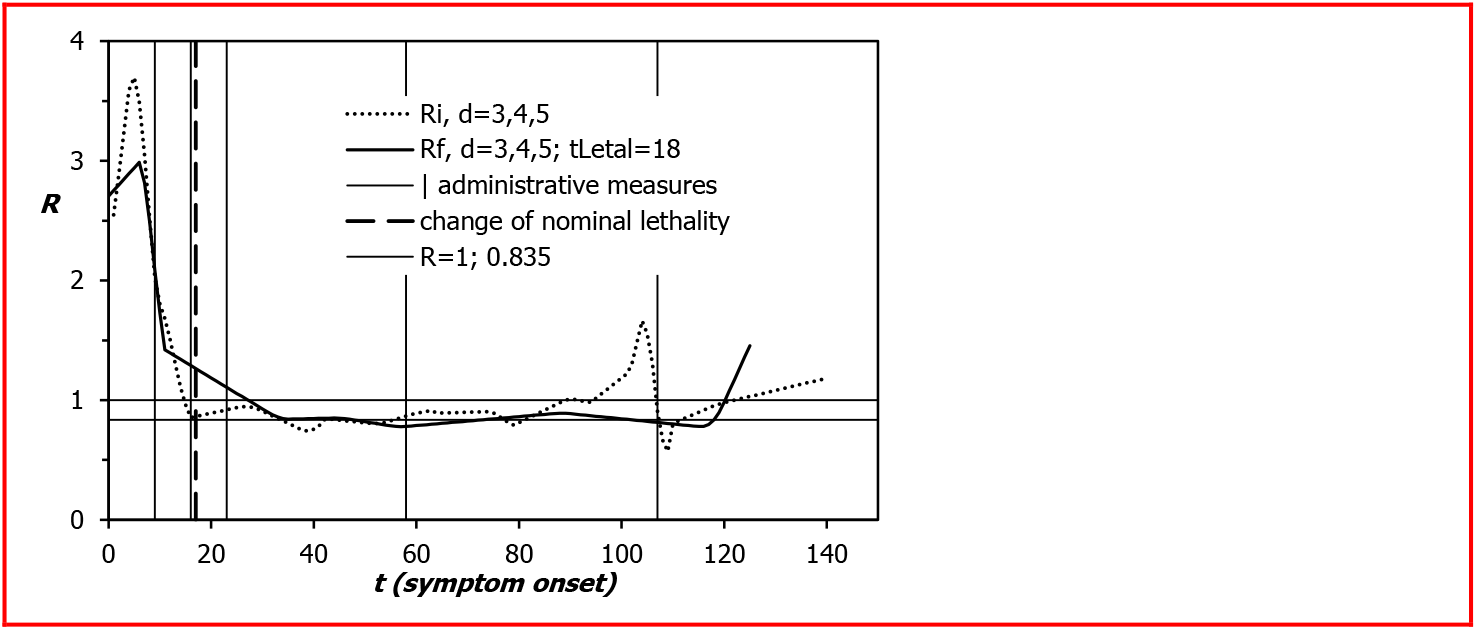
Effective reproduction rates obtained from the reported number of daily infections (*R*_eff_) and from the reported number of fatalities (*R*_f_); change of lethality taken from Figure 5

The coincidence of the essential features of the two curves confirms that the assumed average time between symptom onset and death, *t*_Lethal_ = 18 d, is consistent with the data. *R* = 1 and the long-term effective reproduction rate *R* = 0.835 deduced from Figure 4 are marked by horizontal lines.

Despite differences in detail, there is a remarkable similarity in the main characteristics:

- rapid decrease towards 1, before administrative measures to restrict public and private life are in effect;
- effective reproduction rate about 0.835 in the long run.

The overshoot of *R*_i_ around *t* = 107 not reproduced in *R*_f_ is a statistical artefact due to a superspreading effect (Tönnies 2020) leading to a peak in the reported number of daily infections.

The similarity of the effective reproduction rates *R*_I_(*t*) from infections and *R*_f_(*t*) from fatalities suggests that the dynamics of the infection process may well be captured by analyzing the infection rates under certain circumstances although the number of reported infections is generally considered to be not representative of the infection demographics. However, if it is biased consistently and the infection demographics remains the same in a time span, the effective reproduction rate calculated from the infection trajectory can describe the infection dynamics sufficiently well. A change of the testing procedure or an isolated outbreak leads only to a temporary disturbance.

Administrative lockdown measures do not seem to have an immediate positive effect on the infection dynamics, but the ease of restrictions at *t* = 107 arguably a negative one.

## 5 Conclusions

A trend line *n*_Fcalc_ through the Covid-19 fatality rate can sensibly be modelled similar to the trend line n_Icalc_ for the infection rate, with a seed point representing the strength and a day-to-day reproduction rate representing the dynamics. The effective reproduction rates calculated from the two trend lines are similar if *n*_Fcalc_ is shifted by 18 days, the average time from symptom onset to death, towards earlier times. Its long-term value is around 0.835.

The coincidence of the two reproduction rates indicates that the effective reproduction rate calculated from the number of daily infections is representative of the infection process even if the testing procedure does not capture all cases. It is sufficient that testing bias is consistent and the infection demographics remains the same over a period of time. This seems to be the case for the data reported by the Robert-Koch-Institut.

There is no evidence that administrative measures to contain the epidemic decreases the infection dynamics represented by the reproduction rate of the daily infections. This does not necessarily mean that social distancing is useless but arguably indicates that self-regulatory processes are at least as important as administrative measures. Caveat: absence of evidence is not evidence of absence! Changes in the testing process may blur effects.

The fatality rate can be projected from the infection rate by assuming an average delay of 18 or of 20 days between symptom onset and death, and a time-dependent nominal lethality. A sharp increase of the time-dependent nominal lethality after March 16 (when public life was restricted) from 2.5% to 7% may indicate a change in the infection process with probably more vulnerable people infected relative to the mobile young population infected in leisure-time superspreading events.

The time-dependent nominal lethality decreases from 7% at April 27 to 1% at begin July. Face mask wearing in public places became obligatory at April 27 and seem to have lead to less infections with lethal outcome. Furthermore, officially reported cases in May and June may be less lethal than before. More detailed analyses and epidemiological expertise are necessary to decide on these issues.

## Data Availability

I have used publicly available data from the Robert-Koch-Institut, Berlin, Germany

https://www.rki.de/DE/Content/InfAZ/N/Neuartiges_Coronavirus/Projekte_RKI/Nowcasting_Zahlen.html

https://www.rki.de/DE/Content/InfAZ/N/Neuartiges_Coronavirus/Fallzahlen.html

## References

(Bendavid E and others 2020): Eran Bendavid, Bianca Mulaney, Neeraj Sood, Soleil Shah, Emilia Ling, Rebecca Bromley-Dulfano, Cara Lai, Zoe Weissberg, Rodrigo Saavedra-Walker, James Tedrow, Dona Tversky, Andrew Bogan, Thomas Kupiec, Daniel Eichner, Ribhav Gupta, John Ioannidis, Jay Bhattacharya. COVID-19 Antibody Seroprevalence in Santa Clara County, California. doi: https://doi.org/10.1101/2020.04.14.20062463, posted April 30, 2020 on medRxiv, https://www.medrxiv.org/content/10.1101/2020.04.14.20062463v2

(Böhmer M and others 2020) Böhmer MM, Buchholz U, Corman VM, Hoch M, Katz K, Marosevic DV, Böhm S, Woudenberg T, Ackermann A, Konrad R, Eberle U, Treis B, Dangel A, Bengs K, Fingerle V, Berger A, Hörmansdorfer S, Ippisch S, Wicklein B, Grahl A, Pörtner K, Muller N, Zeitlmann N, Boender TS, Cai W, Reich A, an der Heiden M, Rexroth U, Hamouda O, Schneider J, Veith T, Mühlemann B, Wölfel R, Antwerpen M, Walter M, Protzer U, Liebl B, Haas W, Sing A. Drosten C, Zapf A. Investigation of a COVID-19 outbreak in Germany resulting from a single travel-associated primary case: a case series. The Lancet Infectious Diseases VOLUME 20, ISSUE 8, P920-928, AUGUST 01, 2020. Published: May 15, 2020, DOI:https://doi.org/10.1016/S1473-3099(20)30314-5

(Mergel D 2017) Mergel D. Physik mit Excel und Visual Basic. Springer Spektrum 2017, ISBN 978-3642-37856-0. Chapter 9, Anpassung von Trendkurven an Messpunkte (Fitting trend curves to measurement data).

(Mergel D 2020, 1) Mergel D. Modelling daily infections with Covid-19 in Germany, France, and Sweden with a trend line based on day-to-day reproduction rates. doi: https://doi.org/10.1101/2020.06.03.20121459, posted June 05, 2020 on medRxiv https://www.medrxiv.org/content/10.1101/2020.06.03.20121459v1

(Mergel D 2020, 2) Mergel, D. Covid-19 Fatality Rate Between 0.1 and 0.3%, Gangelt and Santa Clara Combined. Preprints 2020, 2020070155 doi:https://doi.org/10.20944/preprints202007.0155.v1 Version 1 : Received: 6 July 2020 / Approved: 8 July 2020 / Online: 8 July 2020 (12:04:50 CEST)

(RKI 23-07-2020, 1] Robert-Koch-Institut, Tabelle mit Nowcasting-Zahlen zur R-Schätzung, Punktschätzer der Anzahl Neuerkrankungen (ohne Glättung), https://www.rki.de/DE/Content/InfAZ/N/Neuartiges_Coronavirus/Projekte_RKI/Nowcasting Zahlen. html.also: https://en.wikipedia.org/wiki/Template:COVID-19 pandemic data

[RKI 23-07-2020, 2] Robert-Koch-Institut, Todesfälle, https://www.rki.de/DE/Content/InfAZ/N/NeuartigesCoronavirus/Fallzahlen.html, also: https://www.google.com/search?q=covid-19+tote+in+deutschland&rlz=1C1AKJH_enDE808DE831&oq=Covid-19+&aqs=chrome.3.69i57j0j69i59l2j0l4.8335j0j8&sourceid=chrome&ie=UTF-8

(Streeck H and others 2020): Hendrik Streeck, Bianca Schulte, Beate Kuemmerer, Enrico Richter, Tobias Hoeller, Christine Fuhrmann, Eva Bartok, Ramona Dolscheid, Moritz Berger, Lukas Wessendorf, Monika Eschbach-Bludau, Angelika Kellings, Astrid Schwaiger, Martin Coenen, Per Hoffmann, Markus Noethen, Anna-Maria Eis-Huebinger, Martin Exner, Ricarda Schmithausen, Matthias Schmid, Gunther Hartmann. Infection fatality rate of SARS-CoV-2 infection in a German community with a super-spreading event, doi: https://doi.org/10.1101/2020.05.04.20090076 posted June 02, 2020 on medRxiv, https://www.medrxiv.org/content/10.1101/2020.05.04.20090076v2

(Tönnies H 2020) Tönnies Holding. “On Wednesday, 17 June 2020, work was ramped-down at the Tönnies slaughterhouse in Rheda-Wiedenbrück due to an outbreak of COVID-19 infections originating from there. By that date, 657 employees had tested positive for the virus“. https://en.wikipedia.org/wiki/T%C3%B6nnies_Holding, also: https://www.dw.com/en/coronavirus-over-600-people-test-positive-at-german-slaughterhouse/a-53846038

(Verity R and others 2020) Verity R, Okell, LC, Dorigatti I, Winskill P, Whittaker C, Imai N, Cuomo-Dannenburg G, Thompson H, Walker PGT, Fu H, Dighe A, Griffin JT, Baguelin M, Bhatia S, Boonyasiri A, Cori A, Cucunubá Z, FitzJohn R, Gaythorpe K, Green W, Hamlet A, Hinsley W, Laydon D, Nedjati-Gilani G, Riley S, van Elsland S, Volz E, Wang H, Wang Y, Xi X, Donnelly CA, Ghani AC, Ferguson NM. The Lancet Infectious Diseases, VOLUME 20, ISSUE 6, P669-677, JUNE 01, 2020, Estimates of the severity of coronavirus disease 2019: a model-based analysis. Open Access Published: March 30, 2020 DOI:https://doi.org/10.1016/S1473-3099(20)30243-7, https://www.thelancet.com/journals/laninf/article/PIIS1473-3099(20)30243-7/fulltext

